# Increased variability in visual processing in children with autism

**DOI:** 10.64898/2026.10.31.26366985

**Authors:** Brooke Keough, Winko W. An, Virginia Rosenberger, Janani C. Elumalai, Meagan Tsou, Ariana Mechem, Noah Crane, April R. Levin, Carol L. Wilkinson, Charles A. Nelson

**Author notes:** **Corresponding authors:** Charles A. Nelson, PhD, Address: BC525.10, 2 Brookline Place, Brookline, MA USA 02445, Carol L. Wilkinson, MD PhD, Address: BC525.6, 2 Brookline Place, Brookline, MA USA 02445. These authors contributed equally to this work (Brooke Keough, Winko W. An). These authors contributed equally as joint last authors (Carol L. Wilkinson, Charles A. Nelson).

## Abstract

The visual evoked potential (VEP) indexes sensory processing, but the neurophysiological basis of altered VEP amplitudes in autism remains unclear. We examined whether reduced VEP amplitude reflects increased trial-to-trial neural variability and whether this variability relates to electrophysiological markers of cortical excitation/inhibition and developmental functioning. Pattern-reversal VEPs were analyzed in 102 typically developing (TD; 44 female) and 105 autistic (AUT; 15 female) children aged 2–12 years. N1 and N1-P1 amplitudes and inter-trial phase coherence (ITPC) were computed. Resting-state periodic alpha activity and aperiodic exponent were derived using spectral parameterization. Associations among VEP, resting EEG, age, and developmental/behavioral measures were assessed using multiple linear regression. AUT children exhibited reduced N1 amplitude and lower ITPC, with marginally reduced N1-P1 amplitude. Higher ITPC was strongly associated with larger N1 and N1-P1 amplitudes, and group differences in VEP amplitudes were no longer significant after accounting for ITPC. Greater periodic alpha activity was associated with higher ITPC and larger N1-P1 amplitude, while a steeper aperiodic exponent was associated with higher ITPC in the pooled cohort. Within AUT children, higher ITPC was associated with better nonverbal development, verbal cognition, and language scores. Reduced VEP amplitudes in autism may primarily reflect increased trial-to-trial variability rather than systematically weaker stimulus-evoked responses. Associations between ITPC and resting EEG measures suggest that cortical excitation/inhibition may contribute to sensory response reliability. These findings identify neural response consistency as a developmentally relevant feature of visual processing in autism.

## 1. Introduction

Autism spectrum disorder (ASD) is a neurodevelopmental condition defined by deficits in social communication and restricted or repetitive behaviors and interests (*Diagnostic and Statistical Manual of Mental Disorders: DSM-5.*, 2013), with marked heterogeneity in presentations and symptoms across individuals (Amaral et al., 2008; Baiano & Zeppieri, 2023; Halder et al., 2024). Sensory hypo- and hyper-sensitivities are a common feature of autism (Balasco et al., 2020; Takach et al., 2025). Studies examining sensory processing differences have reported that up to 95% of children with autism spectrum disorder exhibit sensory processing dysfunction (Tomchek & Dunn, 2007). The relationship between increased sensory symptoms and elevated social and communication impairment in autism has also been explored (Derakhshanrad et al., 2024; Leekam et al., 2007). Indeed, dysregulation in sensory processing is hypothesized to contribute to social symptoms of autism (Markram & Markram, 2010; Thye et al., 2018), as altered processing of and responsiveness to one’s external environment may affect social responsiveness. Understanding the neural underpinnings of sensory atypicalities in autism may therefore shed light on the developmental pathways contributing to social and behavioral differences and can help identify brain-based markers associated with heterogeneity in this population.

Neuroimaging tools and experimental paradigms have been developed to characterize neural responses to sensory input in autism, among which the visual evoked potential (VEP) has emerged as a promising approach. Using electroencephalography (EEG) and simple visual stimuli, the VEP paradigm was originally developed to investigate the function of the visual pathway from the optic nerve to the visual cortex (Shigihara et al., 2016; Šuštar Habjan et al., 2025). VEPs are typically studied by averaging the evoked response across trials and examining components such as the N1 and P1, which are extracted from the average waveform. The amplitude and latency of these components provide information about neural responses and timing of information transmission through the visual system (Baiano & Zeppieri, 2023; Creel, 2019). In neurotypical adults, VEP measures have shown high reliability across and within sessions (Tello et al., 2010; Trevino et al., 2018). More recently, VEPs have become an increasingly utilized tool for studying atypical visual system development in young children with neurodevelopmental disorders, as they can be reliably elicited using simple visual stimuli that are well tolerated across a wide range of developmental abilities (Köster et al., 2023). However, studies of VEPs in idiopathic autism have reported inconsistent findings, particularly for VEP amplitudes. Decreased N1 (N75) and/or P1 (P100) amplitudes have been reported in some studies (Kovarski et al., 2019; Siper et al., 2016), whereas others found no significant differences (Hamdan et al., 2022; Milne, 2011; Sayorwan et al., 2018; Webb et al., 2023). Similarly, increased, decreased, or unchanged VEP amplitudes have been reported in genetic disorders associated with autism, including Phelan-McDermid syndrome (Siper et al., 2022), Rett syndrome (LeBlanc et al., 2015), fragile X syndrome (Knoth et al., 2014), and tuberous sclerosis complex (Varcin et al., 2016). These observations suggest the need to move beyond conventional VEP amplitude measures to better understand the underlying neurophysiological mechanisms and heterogeneity of visual processing in autism.

Among studies reporting reduced VEP amplitudes in autism, at least two non-mutually exclusive, neurophysiological processes may account for this finding. One possibility is that stimulus-evoked neural responses are intrinsically weaker, potentially reflecting reduced excitatory drive relative to inhibition (LeBlanc et al., 2015). Another possibility is that reduced VEP amplitudes may reflect increased *trial-to-trial variability* in neural responses, whereby less consistent responses across repeated stimulus presentations attenuate the amplitude of the averaged VEP. These two mechanisms may coexist and jointly contribute to the observed reduction in VEP amplitude. This latter hypothesis has gained support from several studies employing single-trial analyses. Using a small sample, Milne (2011) reported greater trial-to-trial variability in P1 amplitude and latency, accompanied by reduced inter-trial phase coherence, in children with autism, suggesting less consistent synchronization of stimulus-evoked neural activity. Kovarski et al. (2019) further demonstrated that the group-level P100 amplitude reduction in autism disappeared after reconstructing VEPs using only trials with consistent responses. Extending this line of work, Dong et al. (2024) introduced a nonlinear mixed-effects modeling framework for trial-level VEP analysis and, using a large multisite cohort, demonstrated that children with autism exhibit significantly greater intra-individual trial-to-trial variability in P1 latency than neurotypical peers. Beyond VEPs, studies using fMRI and steady-state visual evoked responses have also reported greater trial-to-trial neural variability and reduced response reliability in autism (Dinstein et al., 2012; Haigh et al., 2015; Weinger et al., 2014), leading to the proposal of a broader “neural unreliability” framework. However, two important research gaps remain. First, no study has *directly* quantified the relationship between trial-to-trial variability and VEP amplitude, or *statistically* determined whether group differences in VEP amplitude persist after accounting for such variability. Addressing this question is important for determining whether increased trial-to-trial variability can account for reductions in VEP amplitude when they are observed in autism. Second, no study has examined the relationship between trial-to-trial variability and *developmental outcomes* in children with autism. Determining whether this neural measure is associated with cognitive or behavioral functioning is essential for evaluating its potential as a biomarker for prognosis, stratification, and treatment monitoring in autism.

Another important mechanistic question remains unanswered: what gives rise to the heightened trial-to-trial sensory variability, or “neural noise,” that has been proposed to characterize autism (for a review, see Raul et al., 2024)? One influential hypothesis is that this variability arises, at least partially, from an altered balance between neuronal excitation (E) and inhibition (I) (Rubenstein & Merzenich, 2003). Specifically, reduced inhibitory signaling and/or excessive excitation may increase spontaneous neural activity and impair the temporal synchronization of neuronal populations required to generate consistent sensory responses (Foss-Feig et al., 2017). In support of this hypothesis, converging evidence has implicated abnormalities in γ-aminobutyric acid (GABA)-mediated inhibitory signaling in autism (Robertson et al., 2016; Zhao et al., 2022). However, despite the prominence of this hypothesis, the proposed link between altered E/I balance and increased trial-to-trial neural variability has not been directly evaluated. Although E/I balance cannot be measured non-invasively using EEG, recent advances in spectral parameterization have provided putative electrophysiological markers of cortical excitation and inhibition (Donoghue et al., 2020). This approach decomposes the EEG power spectrum into an aperiodic 1/f component and oscillatory (periodic) activity. Of relevance, the aperiodic exponent has been proposed as an indirect marker of E/I balance (Gao et al., 2017), whereas periodic alpha activity has been broadly associated with cortical inhibition (Chapeton et al., 2019; Klimesch et al., 2007; Mathewson et al., 2011). Together, these two measures provide an opportunity to examine whether EEG features proposed to index aspects of cortical excitation and inhibition are associated with inter-trial neural variability in autism.

The present study sought to address these important questions using VEP and resting-state EEG recordings acquired from the same individuals. The study included a large cohort of preschool- and school-aged autistic (AUT) and typically developing (TD) children. From the VEP recordings, we quantified N1 and P1 amplitudes and inter-trial neural variability using inter-trial phase coherence (ITPC). From resting-state EEG, we derived the aperiodic exponent and periodic alpha activity using spectral parameterization. We first characterized group differences in these neurophysiological measures and then examined their associations with one another, as well as with developmental outcomes, including verbal and non-verbal abilities. Together, these analyses aimed to determine whether inter-trial neural variability accounts for altered VEP amplitudes in autism and whether putative electrophysiological markers of cortical excitation and inhibition are associated with this variability. These analyses may provide mechanistic insights into sensory processing abnormalities and their relevance to early neurodevelopment in autism.

## 2. Methods

### 2.1 Participants

Participants were recruited from four studies at Boston Children’s Hospital: Sensory Processing in Neurodevelopment (Study 1; IRB-P00045936), Brain Indicators of Developmental Growth (Study 2; IRB-P00034676), Fragile X Neural Markers (Study 3; IRB-P00025493), and Infant-Sibling Project (Study 4; IRB-P00018377).

A total of 114 typically developing (TD) participants (24–155 months) were recruited across Studies 1, 2, and 4 (Table 1). From the larger pool of TD participants across these studies, participants were selected to age-match the autism cohort. Exclusion criteria for all studies included children born premature or of low birth weight, or children with a known genetic, developmental, or neurological disorder.

**Table 1.** Sample characteristics.

|  | TD | AUT | p value |
|---|---|---|---|
| **Enrollment** |  |  |  |
| Study 1 | 74 | 76 |  |
| Study 2 | 35 | 28 |  |
| Study 3 | 0 | 9 |  |
| Study 4 | 5 | 8 |  |
| *Total enrollment* | 114 | 121 |  |
| **Exclusion** |  |  |  |
| Visual abnormality | 3 | 4 |  |
| Developmental concern | 3 | 2 |  |
| Data quality | 6 | 10 |  |
| **Final samples for analysis\*** | **102** | **105** |  |
| **Female (%)** | 44 (43.1%) | 15 (14.4%) |  |
| **Age in months** |  |  |  |
| Mean (std) | 72.6 (35.4) | 72.6 (33.1) | 1.0 |
| Range | 23.5-155.0 | 24.0-144.0 |  |
| **Race and Ethnicity (%)** |  |  |  |
| American Indian or Alaska Native | 0 (0.0%) | 4 (3.8%) |  |
| Asian | 15 (14.7%) | 4 (3.8%) |  |
| Black or African American | 1 (1.0%) | 9 (8.6%) |  |
| Hispanic or Latino | 2 (2.0%) | 8 (7.6%) |  |
| Middle Eastern or North African | 2 (2.0%) | 1 (1.0%) |  |
| Native Hawaiian or Pacific Islander | 0 (0.0%) | 0 (0.0%) |  |
| White | 58 (56.9%) | 48 (45.7%) |  |
| More Than 1 Race | 24 (23.5%) | 25 (23.8%) |  |
| Unknown/Other | 0 (0.0%) | 6 (5.7%) |  |
| **Income** |  |  |  |
| Less than \$10,000 | 2 (2.0%) | 2 (1.9%) |  |
| Between $10,000 - $40,000 | 1 (1.0%) | 9 (8.6%) |  |
| Between $40,000 - $70,000 | 0 (0.0%) | 18 (17.1%) |  |
| Between $70,000 - $100,000 | 10 (9.8%) | 16 (15.2%) |  |
| Between $100,000 - $140,000 | 11 (10.8%) | 10 (9.5%) |  |
| Greater than \$140,000 | 73 (71.6%) | 38 (36.2%) |  |
| Unknown/Did not Answer | 5 (4.9%) | 12 (11.4%) |  |
| **MSEL: Nonverbal Domain\*\*** |  |  |  |
| n | 30 | 39 |  |
| Mean (range) | 98.2<br>(66.3 - 135.3) | 69.9<br>(53.5 - 109.0) | 4.1e-08 |
| **Vineland: Adaptive Behavior Composite\*\*** |  |  |  |
| n | 28 | 44 |  |
| Mean (range) | 102.2<br>(83.0 - 135.0) | 72.3<br>(43.0 – 108.0) | 3.6e-17 |
| **Preschool Language Scales (PLS)\*\*** |  |  |  |
| n | 28 | 33 |  |
| Mean (range) | 113.8<br>(95.0 – 135.0) | 71.5<br>(50.0 – 116.0) | 6.4e-16 |
| **SRS T-score\*\*\*** |  |  |  |
| n | 98 | 86 |  |
| Mean (range) | 45.6<br>(37.0 - 60.0) | 71.8<br>(47.0 – 91.0) | 1.1e-41 |

\*The remainder of the table contains only subjects within ‘Final samples for analysis’.
\*\*Standard scores reported have a mean of 100 and standard deviation of 15. Scores from assessments conducted concurrently with EEG acquisition are reported.
\*\*\*T-score reported has a mean of 50 and standard deviation of 10; scores under 60 are within normal limits.

A total of 121 participants with autism spectrum disorder (ASD) (24–144 months) were recruited across the four studies (Table 1). In Studies 1–3, a community diagnosis of ASD was required. Studies 2 and 3 recruited participants with autism (AUT) in order to match a Fragile X cohort, and thus confirming the autism diagnosis was required via Social Responsiveness Scale 2 (SRS-2, Study 2) and the Autism Diagnostic Observation Schedule-2 (ADOS-2, Study 3). As Study 2 was investigating language, a score of below 85 on either the auditory comprehension or expressive communication subscale of the Preschool Language Scales was also required for inclusion. In Study 4, the diagnosis of ASD was established via the ADOS. Exclusion criteria for all studies included children born premature or of low birth weight, or children with a known genetic disorder.

Twelve typically developing children and 16 children with autism were excluded from analysis due to insufficient EEG data defined by fewer than 40 trials (n=16), ineligibility determined post-study for developmental (n=5), or uncorrected visual abnormalities (n=7). Total samples for final analysis included 102 typically-developing and 105 autistic children.

### 2.2 Developmental and Behavioral Measures

Developmental and behavioral data were collected as part of the research protocol in Studies 2–4 or obtained through post hoc medical record review in Study 1. Measures were harmonized across studies into four domains: Language, Verbal Cognition, Nonverbal Cognition, and Adaptive Behavior. Age-normed standard scores (mean = 100, SD = 15) were used for all domains. Assessments in Studies 2–4 were collected concurrently with EEG, whereas assessments identified through medical record review in Study 1 were included based on prespecified criteria regarding their timing relative to EEG acquisition. Detailed assessment, harmonization, and inclusion procedures are provided in the Supplemental Information Section 1.

### 2.3 Experiment and EEG Collection

EEG data were acquired during resting-state and visual evoked potential (VEP) paradigms. Here, resting-state EEG was operationally defined as EEG recorded in the absence of time-locked stimuli, as children in this age range and with varying developmental abilities were often unable to reliably follow instructions to remain at rest (Webb et al., 2020). During resting-state EEG, participants watched a screen-saver video or a preferred video for 3–5 minutes while instructed to remain quiet and still. For the VEP paradigm, pattern-reversal black-and-white checkerboards were presented at 2 Hz on a 60-Hz monitor using E-Prime (v3, Psychology Software Tools, Inc.). Each checkerboard square subtended a visual angle of approximately 2°. Studies 1 and 2 continuously presented 400 and 200 trials, respectively, whereas Studies 3 and 4 used Tobii Eye-Tracking to present 200 and 100 gaze-dependent trials, respectively, when participants were looking at the screen. EEG was recorded in a dimly lit, sound-attenuated, electrically shielded room. Participants sat independently or on a caregiver’s lap, with caregivers instructed not to speak or socially engage with the child during recording.

EEG data were acquired using a 128-channel HydroCel Geodesic Sensor Net (Version 1, EGI Inc., Eugene, OR) connected to a DC-coupled amplifier (Net Amps 400, EGI Inc., Eugene, OR). In Studies 1-3, electrode impedances were maintained below 50 kΩ, and data were sampled at 1,000 Hz. In Study 4, electrode impedances were maintained below 100 kΩ, and the sampling rate was set at 500 Hz. All studies used Cz as the reference electrode.

### 2.4 EEG Preprocessing

Following acquisition, session videos were reviewed using AVOCODO (Audio/VideO CODing Optimization) (An, 2026), a MATLAB-based behavioral coding tool. Periods in which the participant was looking away from the screen or had significant behavioral disruptions such as crying were annotated and corresponding trials excluded from analysis. EEG data were batch-processed in MATLAB using the Harvard Automated Preprocessing Pipeline for EEG (HAPPE, v3; Gabard-Durnam et al., 2018; Lopez et al., 2022). Processing included CleanLine line-noise removal, resampling to 250 Hz, 100-Hz low-pass filtering, detection and interpolation of bad channels, and wavelet-thresholding artifact removal (Castellanos & Makarov, 2006). Data were subsequently bandpass filtered at 1–100 Hz for resting EEG and 0.3–30 Hz for VEP. Resting EEG was segmented into 2-s epochs with 50% overlap. VEP data were epoched from −100 to 400 ms relative to stimulus onset and baseline corrected using the −100 to 0 ms interval. For both VEP and resting EEG, epochs containing amplitudes exceeding ±200 µV were excluded from analysis. Participants with fewer than 40 usable VEP trials were excluded, yielding 102 TD and 105 AUT participants (Table 1). To account for differences in trial number, 40 trials were randomly selected from each participant for analysis.

### 2.5 Visual Evoked Potential (VEP) Analysis

#### 2.5.1 PCA-based Electrode Selection

Because the spatial distribution of VEP responses and precise electrode positioning may vary across participants, we used a data-driven approach to select VEP electrodes individually rather than relying on the conventional O1, Oz, and O2 electrodes. For each participant, spatial principal component analysis (PCA) was applied to trial-averaged VEPs within a predefined posterior electrode region, and the three electrodes contributing most strongly to the dominant spatial patterns were selected for analysis. PCA was used only for electrode selection; VEP measures were subsequently derived from the original EEG signals at the selected electrodes. Detailed procedures and comparison with conventional electrode selection are provided in the Supplemental Information (Section 2; Figures S2–S3).

#### 2.5.2 VEP components and amplitude calculations

We focused on two primary components of the VEP waveform, N75 (N1) and P100 (P1) (Šuštar Habjan et al., 2025). The later N2/N145 component was not examined because it was less consistently identifiable across participants than the earlier N1 and P1 components. Notably, prior work has reported a less well-defined N145 with larger (1°) than smaller (0.25°) checks (Šuštar Habjan et al., 2025); the present study used an even larger check size (2°). We defined the N1 and P1 components and calculated their amplitude as follows.

The P1 component was defined as the first peak closest to 110 milliseconds (ms); this starting point was chosen based on the mean latency for all subjects in the study. P1 latency was calculated as the distance from stimulus onset (0 ms) to this peak, occurring within the range of 84 to 188 ms for all subjects. A participant was determined to have a “double peak” if, within this range, a second peak greater than half the amplitude of the first peak was found (LeBlanc et al., 2015). In this case, N1 was defined as the first minimum peak preceding the P1, occurring within the range of 40 to 88 ms for all participants.

The absolute amplitude (from 0 µV) for N1 and P1 was calculated at the identified N1 and P1 peaks. We also extracted the N1-P1 amplitude, defined as the amplitude difference between absolute P1 and N1 amplitudes (Šuštar Habjan et al., 2025).

#### 2.5.3 Inter-trial phase coherence (ITPC)

Trial-to-trial variability in VEP signals was quantified using inter-trial phase coherence (ITPC), calculated with the following equation (An et al., 2022):

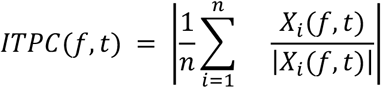

where Xi(f, t) denotes the Short-Time Fourier Transform (STFT) of a given time course x(t) at frequency f, and n denotes the number of trials in a condition. The value of ITPC ranges between 0 and 1, with 0 indicating totally random phase distribution and 1 indicating perfect phase synchronization across trials.

To derive a summary ITPC measure, we applied a cluster-based permutation test (Maris & Oostenveld, 2007) to identify time-frequency regions differing between the TD and AUT groups. ITPC was first averaged across channels for each participant. Clusters were defined using an F-statistic threshold corresponding to p<0.01, and significance was determined from 10,000 permutations of group labels (p_cluster<0.01). ITPC was then averaged within the resulting significant cluster mask for each participant. This summary measure was used in subsequent VEP, resting EEG, and developmental/behavioral analyses, but not to independently test group differences because the mask was defined by the group contrast.

### 2.6 Spectral Parameterization in Resting EEG

Power spectral density (PSD) was estimated using Welch’s method with a Hamming window. Aperiodic power components were estimated using the specparam Python package (Donoghue et al., 2020). This method fits a 1/f line in the log(power)-log(frequency) space across 2-45 Hz. Hyperparameters of this model include aperiodic_mode = ‘fixed’, peak_width_limits = [1,12], max_no_peaks = 8, and peak_threshold = 2 (Hill et al., 2022). Participants with poor model fit, defined as R^2^<0.90, were excluded, resulting in 87 AUT and 99 TD participants. Among retained participants, mean R^2^ was 0.978±0.023 (range: 0.905–0.997) in the AUT group and 0.985±0.018 (range: 0.913–0.994) in the TD group.

The aperiodic exponent was defined as the negative slope of the fitted 1/f line in log-log space. Periodic activity is defined as the residual spectral power after removal of the 1/f estimated aperiodic component. We then defined periodic alpha activity as the average periodic activity between 6-13 Hz (Hill et al., 2022; Wilkinson et al., 2024). Both aperiodic and periodic measures were averaged across three electrodes for comparison, O1, Oz, and O2.

### 2.7 Statistical Analysis

We applied multiple linear regression using statsmodels (v0.14.6) to assess associations between (1) VEP measures and age, (2) VEP amplitudes and ITPC, and (3) VEP and resting EEG measures. Models accounted for group, age, sex, and study, as applicable. For each association, we additionally tested the interaction between groups to determine whether the relationship differed between AUT and TD children. VEP-resting EEG associations were also examined without adjustment for groups to assess relationships across the full cohort irrespective of autism diagnosis.

Finally, within autistic children, we assessed associations between VEP measures and developmental and behavioral outcomes, accounting for age, sex, study, and the time interval between EEG acquisition and the corresponding outcome assessment.

**Figure 1.**
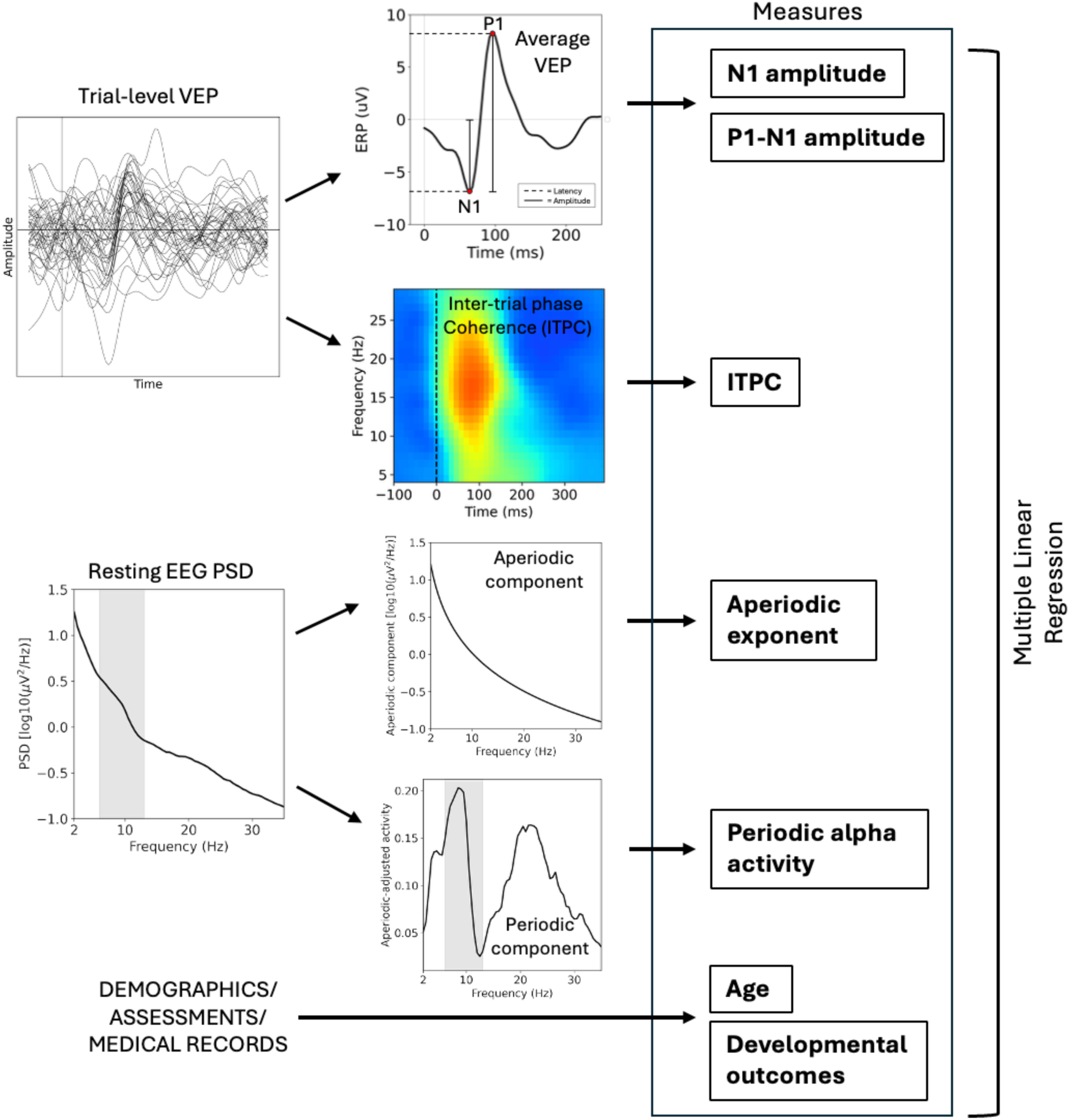
A schematic of the study. Raw visual evoked potential (VEP) and resting EEG data were preprocessed using HAPPE, an automated preprocessing pipeline. From VEP, N1 and N1-P1 amplitudes, as well as inter-trial phase coherence measures were extracted. From resting EEG, aperiodic exponent and periodic alpha activity measures were extracted. Multiple linear regressions were performed to assess the relationship among these measures and their association with age and developmental outcomes.

## 3. Results

### 3.1 VEP

A clear VEP waveform, characterized by distinct N1 and P1 components, was observed in both the AUT and TD groups (Figure 2A). No significant group interactions were observed for the associations between VEP amplitudes and age (all p>0.242). After controlling for age, study, and sex, the AUT group exhibited significantly reduced N1 amplitude compared with the TD group (p = 0.011; Figure 2B). In addition, the N1-P1 amplitude showed a trend toward being smaller in the AUT group than in the TD group (p = 0.071; Figure 2C). No significant age effects were observed, though N1 amplitude showed a marginal significance (p=0.085; Figure 2B).

**Figure 2.**
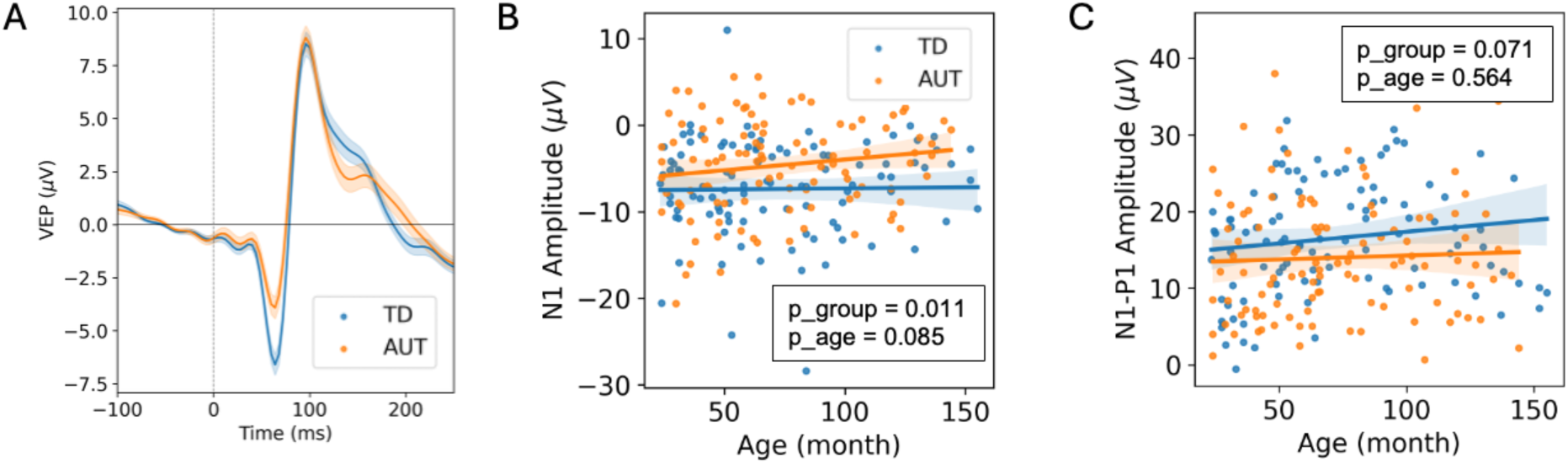
Comparison of VEP waveform and amplitude between AUT and TD groups. A: The average VEP waveforms. B & C: Comparison of N1 and N1-P1 amplitude between groups, respectively, while controlling for age. The p values indicate main effects of age and group.

### 3.2 ITPC

Both the AUT and TD groups exhibited robust ITPC responses, with prominent centroids localized to the N1-P1 complex (∼50–150 ms) and within the canonical alpha–beta frequency range (10–20 Hz) (Figure 3A & 3B). A cluster-based permutation analysis identified a time–frequency cluster in which ITPC was significantly reduced in the AUT group relative to the TD group (p = 0.0004; Figure 3C). Regression analysis confirmed significantly lower mean ITPC extracted from this cluster (hereafter referred to as “mean ITPC”) in the AUT group after controlling for age, sex, and study (p = 0.0003; Figure 3D). There was also a significant main effect of age, with higher ITPC associated with older age (p = 0.005), but no significant age-by-group interaction (p=0.903).

**Figure 3.**
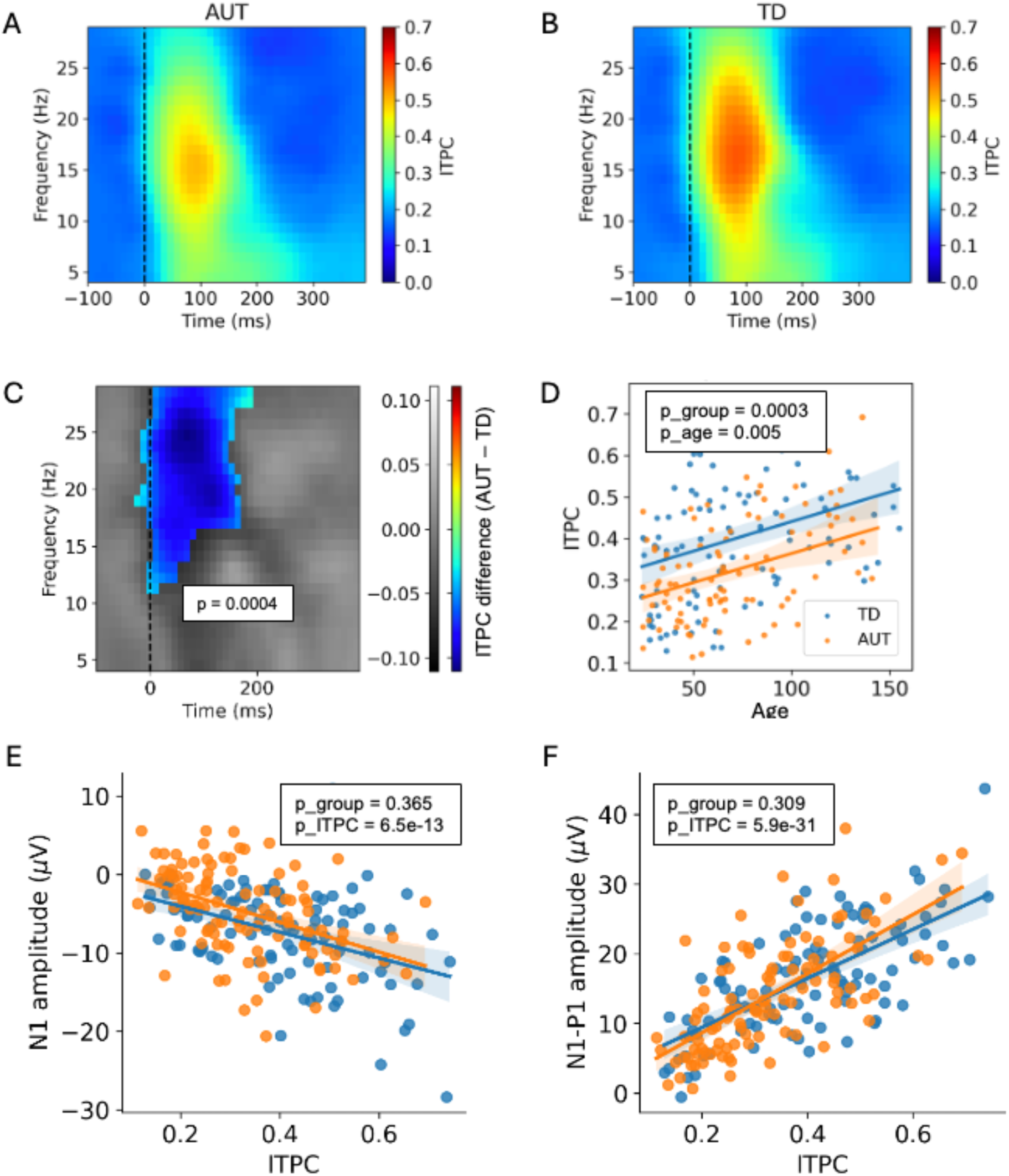
Inter-trial phase coherence (ITPC) differences between autistic (AUT) and typically developing (TD) children and their associations with age and VEP amplitudes. (A–B) ITPC in the AUT (A) and TD (B) groups. (C) ITPC difference between groups (AUT − TD). The colored mask indicates a significant time–frequency cluster identified using a cluster-based permutation test, with nonsignificant regions shown in gray. (D) Association between age and mean ITPC extracted from the significant cluster in (C). (E–F) Associations between mean ITPC and N1 (E) and N1-P1 (F) amplitudes. The p values in (E–F) indicate statistical significance after controlling for group, age, sex, and study.

### 3.3 Association between VEP and ITPC

No significant group interactions were observed for the associations between mean ITPC and VEP amplitudes (all p>0.285). The mean ITPC extracted from the significant time–frequency cluster was strongly associated with greater N1 amplitude (p = 6.5e-13) and the N1-P1 amplitude difference (p = 5.9e-31), after controlling for age, sex, and group (Figure 3E & 3F). When ITPC was included as a covariate, the previously observed group differences in VEP amplitudes were no longer significant (N1: p = 0.227; N1-P1: p = 0.255). These results suggest that individual variability in phase consistency accounts for a substantial proportion of the group-level differences in VEP amplitudes.

### 3.4 Association between VEP and resting EEG measures

A subset of participants contributed usable data from both the VEP and resting-state EEG paradigms (TD: n = 99; AUT: n = 87). In models adjusted for the diagnostic group, higher periodic alpha activity was significantly associated with greater N1-P1 amplitude and higher ITPC (Table S2). No significant group interactions (all p>0.165) were observed for any of these associations, suggesting that the relationships did not significantly differ between AUT and TD groups. When diagnostic group was not included as a covariate (i.e., controlling only for age, study, and sex), all of these associations reached statistical significance, and higher periodic alpha activity was additionally associated with greater N1 amplitude (Table S2; Figure 4).

**Figure 4.**
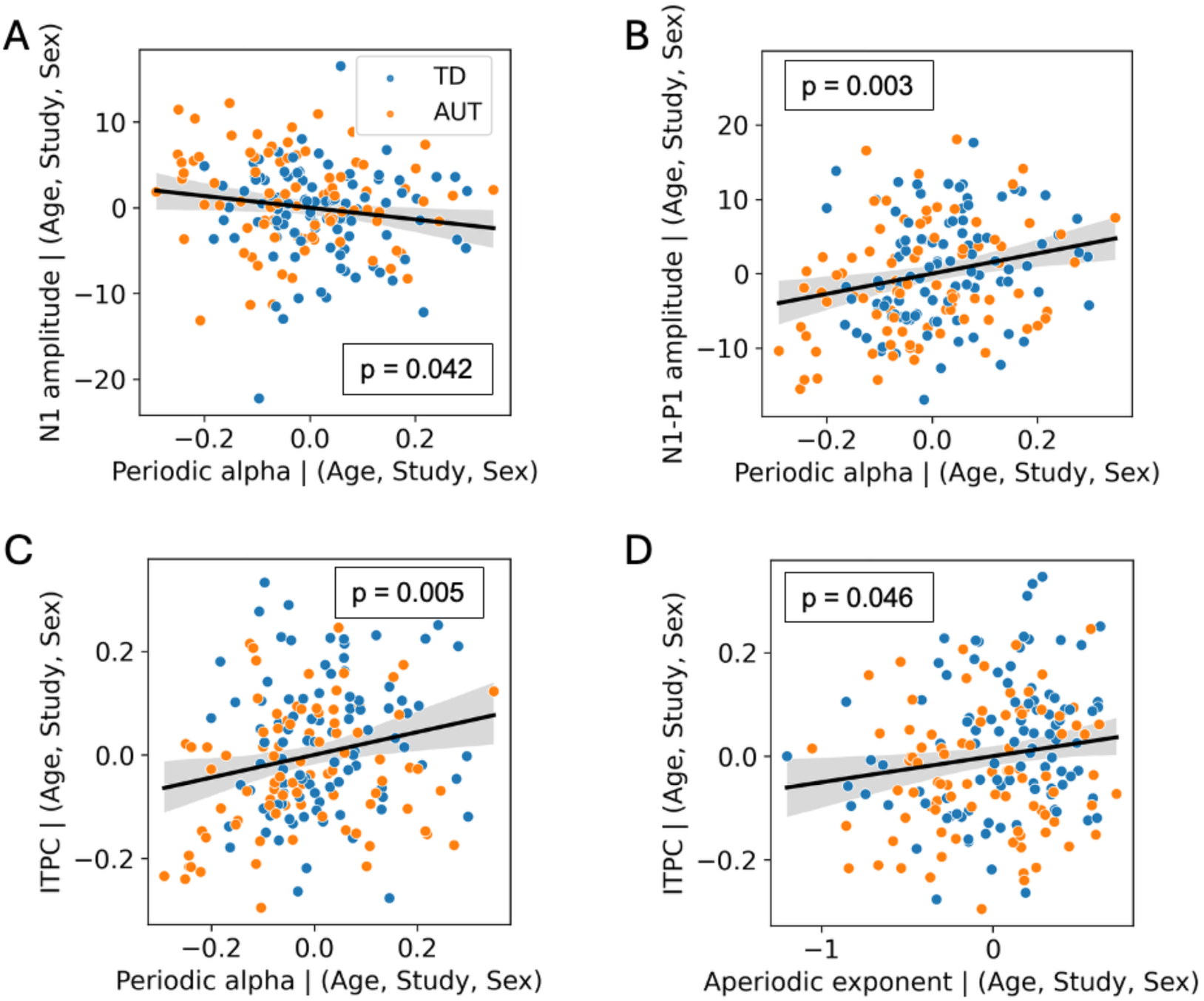
Significant associations between VEP and resting-state EEG measures after controlling for age, sex, and study. Residualized values are plotted to illustrate each association.

### 3.5 Association between VEP and developmental/behavioral measures

No significant associations were observed between N1 or N1-P1 amplitudes and developmental or behavioral measures (Table S3). In contrast, higher ITPC was significantly associated with higher Nonverbal Development (p = 0.0354), Verbal Cognition (p = 0.0473), and Language (p = 0.0295) scores (Table S3; Figure 5).

**Figure 5.**
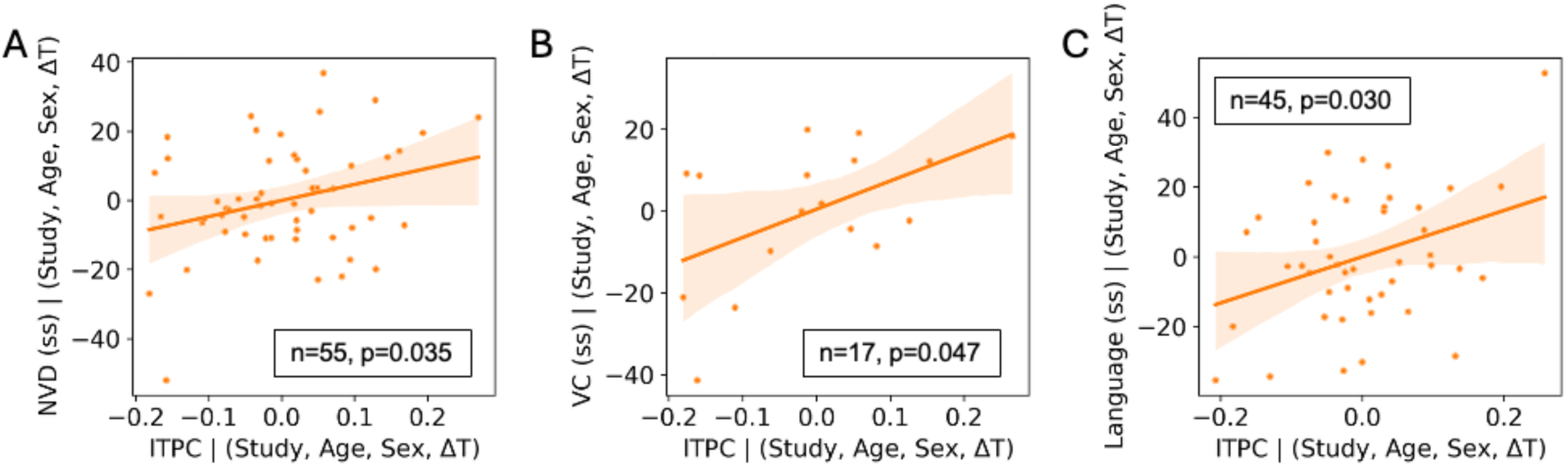
Significant associations between ITPC and the standard score of nonverbal development (NVD), Verbal Cognition (VC), and Language, within the AUT cohort. Residualized values are plotted to illustrate each association.

## 4. Discussion

We recorded visual evoked potentials (VEPs) and resting EEG from children aged 2–12 years with and without autism. We compared VEP component amplitudes across groups and found significantly reduced N1 and marginally reduced N1-P1 amplitudes in the autism (AUT) group. These VEP differences were significantly associated with, and largely explained by, the inter-trial phase coherence (ITPC), a measure of trial-to-trial variability in visual stimulus processing. Moreover, ITPC was positively correlated with the aperiodic exponent and periodic alpha activity in resting EEG, suggesting a potential mechanistic link between greater visual processing variability and reduced network inhibition and/or excitation-inhibition imbalance. Finally, within the autism group, higher nonverbal cognitive and language abilities were significantly associated with greater ITPC, indicating that visual processing consistency may play an important functional role in early cognitive development and learning.

### 4.1 Inter-trial variability largely accounts for reduced VEP amplitudes in autism

Using a large cohort of children, we demonstrated decreased N1 and marginally decreased N1-P1 amplitudes in autism. These findings are broadly consistent with prior studies on children with idiopathic autism (Kovarski et al., 2019; Siper et al., 2016) as well as in some autism-associated neurodevelopmental disorders such as Rett syndrome (LeBlanc et al., 2015; Saby et al., 2023) and Phelan-McDermid syndrome (Siper et al., 2022). Conversely, other autism-associated disorders, such as fragile X syndrome, have reported a higher N1 amplitude compared to controls (Knoth et al., 2014), suggesting that distinct neurobiological mechanisms may underlie sensory processing abnormalities across these genetically defined disorders.

Importantly, we provide direct evidence that these conventional VEP amplitude differences are closely related to trial-to-trial phase consistency, as indexed by inter-trial phase coherence (ITPC), and may not reflect intrinsically weaker single-trial responses. Specifically, autistic children exhibited significantly lower stimulus-evoked phase synchrony than typically developing peers, indicating less consistent neural responses across repeated presentations of identical visual stimuli. Furthermore, the group differences in N1 and N1-P1 amplitudes were no longer significant after accounting for ITPC, suggesting that increased inter-trial variability provides a more compelling explanation for the observed VEP amplitude reductions than intrinsically weaker stimulus-evoked neural responses. Together, these findings are consistent with previous reports of increased inter-trial variability in autistic children (Kovarski et al., 2019; Milne, 2011) and provide direct quantitative support for this proposed mechanism.

To our knowledge, this study is the first to report a significant cross-sectional age-related increase in ITPC in both autistic and typically developing children. Although our cross-sectional data do not allow us to conclude that inter-trial variability decreases within individuals over development, they raise important questions regarding the developmental trajectory of neural response consistency in autism. Sensory experience is known to continuously refine cortical circuits through experience-dependent plasticity, leading to progressively more specialized and efficient sensory processing over development (for review, see Ribic, 2020). Although this process has not been explicitly examined in relation to inter-trial variability in autism, it is possible that experience-dependent refinement also shapes sensory development in autistic individuals, albeit potentially through altered or delayed developmental pathways (Beker et al., 2018; Robertson & Baron-Cohen, 2017). Within this framework, the observed age-related increase in ITPC may reflect progressively more synchronized and reliable neural responses to repeated visual stimulation as cortical circuits mature. Longitudinal studies are needed to determine whether neural response consistency increases within individuals across development and whether this developmental trajectory differs between autistic and typically developing children.

### 4.2 Altered neuronal excitation/inhibition is a plausible mechanism for increased sensory variability in autism

More pronounced periodic alpha activity and a steeper aperiodic exponent during resting-state EEG were both associated with greater ITPC and larger VEP amplitudes. Alpha oscillations have long been associated with cortical inhibition and attentional gating (Chapeton et al., 2019; Klimesch et al., 2007; Mathewson et al., 2011), whereas the aperiodic exponent has been proposed to reflect the balance between cortical excitation and inhibition (E/I), with steeper exponents indicating relatively greater inhibitory tone (Gao et al., 2017; Waschke et al., 2021). Together, these findings suggest that stronger cortical inhibition is associated with more reliable stimulus-evoked neural responses, and provide support for the hypothesis that altered E/I balance contributes to increased neural variability, or “neural noise,” in autism (Raul et al., 2024; Rubenstein & Merzenich, 2003). While previous studies have proposed this relationship conceptually, our findings quantitatively link putative resting-state markers of cortical inhibition to the consistency of sensory-evoked neural responses.

One plausible mechanism is that inhibitory circuits stabilize the recruitment of neuronal populations during sensory processing (Isaacson & Scanziani, 2011). GABAergic inhibition sharpens stimulus selectivity and suppresses competing neural responses, thereby promoting consistent activation of similar neuronal populations across repeated presentations of the same stimulus. Conversely, when inhibitory control is reduced, cortical activity becomes more susceptible to ongoing spontaneous activations, such that identical sensory inputs may recruit partially different neuronal populations or engage them with less precise temporal coordination across trials. This would manifest as reduced phase consistency (lower ITPC) and, consequently, smaller averaged VEP amplitudes due to greater cancellation during trial averaging. This interpretation is consistent with evidence of altered GABAergic signaling in autism (Robertson et al., 2016) and theoretical models proposing that reduced inhibition destabilizes cortical computations and increases intrinsic neural noise.

Several considerations should be noted when interpreting these findings. First, our results should not be interpreted as suggesting that greater inhibition or lower neural noise is invariably beneficial. According to the High Noise-Stochastic Resonance (HNSR) framework (Raul et al., 2024), an intermediate level of intrinsic neural noise may enhance the detection of weak sensory inputs, whereas excessive noise compromises the fidelity and reliability of sensory representations. Therefore, our findings are more consistent with the notion that elevated neural variability becomes maladaptive when it exceeds an optimal range, rather than implying that neural noise should be minimized. Second, we do not propose altered E/I balance to be the sole contributor to the elevated visual processing variability observed in autism. Indeed, the associations between resting EEG measures and ITPC were only moderate, indicating that substantial variance remains unexplained. Other factors, including ongoing fluctuations in cortical state, attention, neuromodulatory processes, and large-scale network organization, may also influence the consistency of sensory processing across repeated stimulus presentations.

### 4.3 Increased visual processing variability is associated with lower developmental scores

Our exploratory analyses further suggested that higher ITPC, reflecting greater consistency in visual processing, was associated with better nonverbal cognition, verbal comprehension, and language abilities. Although derived from a visual paradigm, this finding may reflect a broader property of cortical sensory processing rather than a modality-specific phenomenon. Reliable sensory representations provide a critical foundation for early brain development, as experience-dependent plasticity relies on repeated and consistent sensory inputs to refine cortical circuits, strengthen relevant synaptic connections, and establish efficient neural representations (Hensch, 2005; Ribic, 2020). Such reliable sensory encoding is particularly important during early childhood, when the acquisition of higher-order cognitive functions, including language, depends on accurate and consistent sensory feedback from the environment (Kuhl, 2004, 2010; Werker & Hensch, 2015). Within this framework, increased trial-to-trial variability may reduce the fidelity of sensory representations available for learning, thereby limiting the efficiency with which sensory experiences are translated into cognitive and language development. Although these cross-sectional associations cannot establish causality, they suggest that sensory response consistency may serve as a concurrent marker of developmental functioning in autism and motivate future longitudinal studies to determine whether early neural variability predicts later developmental outcomes.

A recent study of infants with a high familial likelihood for autism reported that greater trial-to-trial variability in VEPs during the first year of life predicted better language and cognitive outcomes at 24 months (Dickinson et al., 2026). These seemingly contradictory findings, however, may instead provide important insight into the changing role of neural variability across development. During infancy, the cortex undergoes rapid expansion of excitatory synaptic connections and exhibits exceptionally high levels of experience-dependent plasticity. Greater neural variability during this period may therefore reflect a more dynamic and adaptable cortical system that facilitates circuit refinement and learning. As development progresses, however, maturation of inhibitory circuitry together with activity-dependent synaptic pruning gradually stabilizes cortical networks, increasing the fidelity and reliability of sensory representations (Hensch, 2005). Within this developmental framework, greater neural variability may be advantageous during infancy by promoting plasticity, whereas lower variability during the preschool and school years may instead reflect more mature and efficient sensory processing. Together, these findings suggest that the functional significance of neural variability is developmentally dependent, with different levels of variability supporting optimal brain function at different stages of neurodevelopment.

### 4.4 Limitations

Several limitations of this study should be acknowledged. First, the cross-sectional design precludes direct characterization of within-individual developmental changes or causal relationships among cortical inhibition, neural variability, and developmental outcomes. Longitudinal studies spanning infancy through childhood will be important for delineating the developmental trajectories of these neurophysiological measures. Second, consistent with the higher prevalence of autism in males, our cohort was predominantly male, which limited our ability to examine potential sex differences in these neural measures. Finally, developmental assessments were available for only a subset of participants, and when available, cognitive testing was not always conducted concurrently with EEG acquisition. These factors likely reduced statistical power and may have attenuated the observed brain–behavior associations. Future studies with more comprehensive and temporally aligned phenotyping will be important for further establishing the relationship between neural response consistency and developmental functioning.

## 5. Conclusion

In a large cohort of autistic and typically developing children, we demonstrate that reduced VEP amplitudes in autism are closely related to increased trial-to-trial variability in visual neural responses. Group differences in VEP amplitudes were no longer significant after accounting for ITPC, suggesting that conventional averaged VEP measures may partly reflect the consistency, rather than simply the strength, of underlying stimulus-evoked responses. Furthermore, associations between ITPC and resting-state periodic alpha activity and aperiodic exponent provide support for a potential contribution of cortical excitation/inhibition to sensory response reliability. Finally, the association between greater ITPC and better cognitive and language abilities in autistic children highlights the potential developmental relevance of neural response consistency. Together, these findings position trial-to-trial neural variability as an important dimension of sensory processing in autism.

## Acknowledgements

We are deeply grateful to all of the children and families who participated in this research and generously contributed their time. We thank the many research assistants, students, and study coordinators whose efforts in participant recruitment and data collection made this work possible. We also extend special thanks to Dr. Rebecca Thomas for her valuable input on the developmental and behavioral measures used in this study.

This work was supported by grants from the Simons Foundation and from NIH (R01-DC010290) to C.A.N; grants from the NIH (1K23DC017983-01A1 and 1T32MH112510), FRAXA Foundation, Autism Science Foundation, The Pierce Family Fragile X Foundation, Thrasher Research Fund, and Harvard Catalyst Medical Research Investigator Training Award to C.L.W.

## Conflict of Interest

The authors declare no conflicts of interest.

## Data Availability

The data that support the findings of this study are available on request from the corresponding author. The data are not publicly available due to privacy or ethical restrictions.

## Code Availability

The code used for VEP, ITPC, resting-state spectral analyses, and statistical analyses is available at the OSF repository: https://osf.io/3w7y2/overview.

## Author Contributions

B.K., W.W.A., A.R.L., C.L.W., and C.A.N. designed research; C.L.W. and C.A.N. acquired funding and resources, and supervised the project; B.K., W.W.A., V.R., J.C.E., M.T., A.M., N.C., and C.L.W. performed investigation and project administration, and curated data; B.K. and W.W.A. conceptualized the project, developed the methodology, created the software, performed formal analysis and visualization; V.R., J.C.E., M.T., A.M., N.C., A.R.L., C.L.W., and C.A.N. reviewed and edited the paper; and B.K. and W.W.A. wrote the paper.

## Notes

### Competing Interest Statement

The authors have declared no competing interest.

### Author Declarations

IRB of Boston Children s Hospital gave ethical approval for this work.

