## Supplemental materials for "Increased variability in visual processing in children with autism"

### Supplementary Information

#### 1. Harmonization of developmental and behavioral measures

Developmental and behavioral measures varied across the four parent studies. Children in Studies 2 and 3 completed the Mullen Scales of Early Learning (MSEL) (Mullen, 1995), Vineland Adaptive Behavior Scales, Third Edition (Vineland-3) (Sparrow et al., 2016), and Preschool Language Scales (PLS) (Zimmerman et al., 2011), whereas children in Study 4 completed the MSEL. These assessments were administered concurrently with EEG acquisition. Children in Study 1 did not complete developmental or behavioral assessments at the time of EEG acquisition; instead, relevant assessments were identified through a post hoc review of available medical records, as described in detail below.

In Study 1, because developmental assessments identified through medical record review were obtained at varying time points relative to EEG acquisition, prespecified timing criteria were applied. Assessments completed at or after 6 years of age were included regardless of their timing relative to EEG acquisition. Assessments completed before 6 years of age were required to have occurred within 12 months of EEG acquisition. These criteria were intended to prioritize assessments temporally proximal to EEG during earlier childhood, while permitting later assessments to characterize developmental outcomes. The distribution of assessment age relative to age at EEG acquisition for each developmental domain is shown in Figure S1.

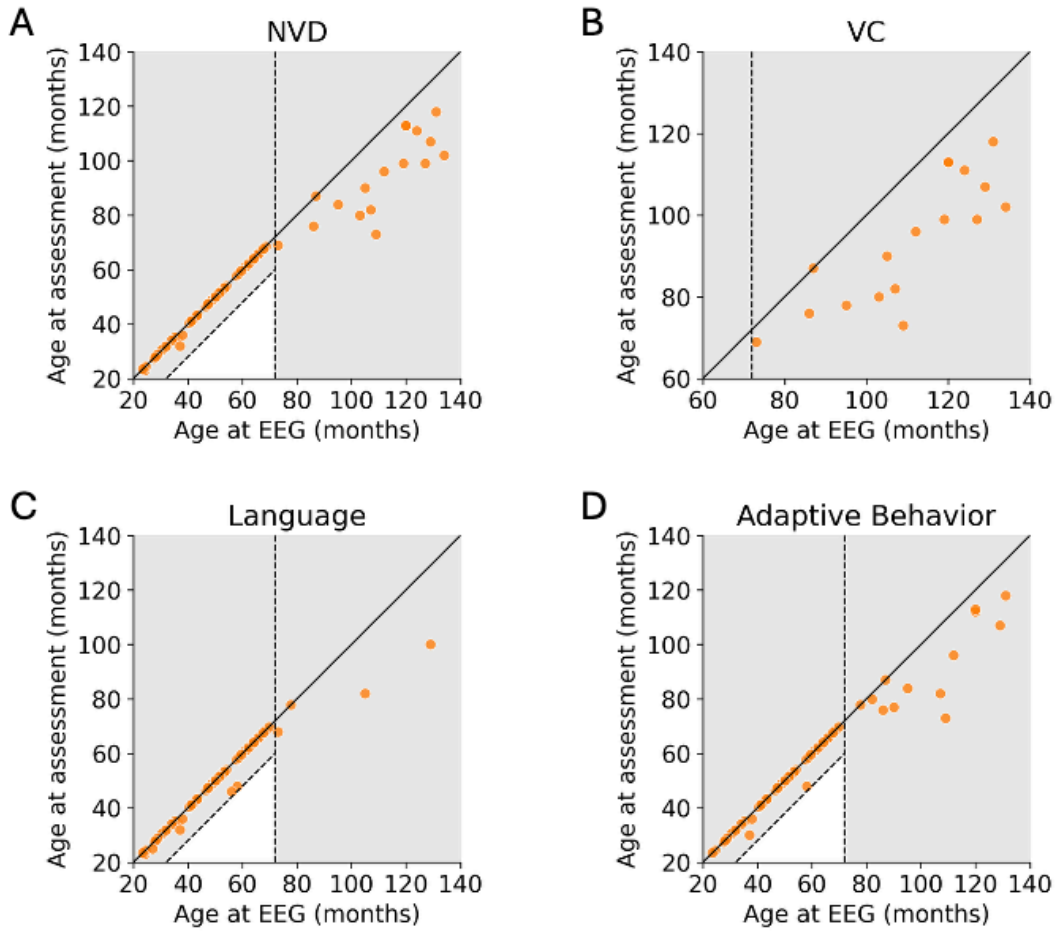


**Figure S1. Timing of developmental assessments relative to EEG acquisition.** Scatterplots show age at EEG acquisition and age at developmental assessment for participants with available measures of (A) nonverbal development (NVD), (B) verbal cognition (VC), (C) language, and (D) adaptive behavior. Each point represents one participant. The solid diagonal line indicates identical ages at EEG and assessment. The dashed diagonal line indicates an assessment occurring 12 months before EEG acquisition, and the dashed vertical line indicates an EEG age of 72 months. Gray shading denotes regions outside the prespecified timing criteria for inclusion in the developmental analyses.

Across all studies, the following variables were filled using the measures as listed. See Table S1 for breakdown of scoring calculations.

Nonverbal cognition was calculated using one of the following: DIfferential Abilities Scale – Second Edition (DAS2) (Elliott, 2007), Stanford-Binet Intelligence Scales – Fifth Edition (SB-5) (Roid, 2003), the Wechsler Intelligence Scales for Children/Wechsler Preschool and Primary Scale of Intelligence (WISC/WPPSI) (Wechsler, 2012, 2014), or the Mullen Scales of Early Learning (MSEL). From each assessment, nonverbal cognition or reasoning standard scores, or combined subcategories meant to together capture nonverbal intelligence, with a mean of 100 and a standard deviation of 15 were extracted. This score captures a child's cognitive abilities without including verbal language abilities as a factor. For the MSEL, the average of relevant subscale T-scores (mean = 50, SD = 10) was calculated. To facilitate comparisons across assessments, the averaged T-scores were linearly rescaled to a standard-score metric (mean = 100, SD = 15) using the following formula:

$$Standard\ Score\ =\ 1.5 \times (Tscore\ -\ 50) + 100$$

These rescaled scores were used for cross-assessment comparisons and should not be interpreted as independently normed composite standard scores.

Verbal cognition was calculated using one of the following: Differential Abilities Scale – Second Edition (DAS2), Stanford-Binet Intelligence Scales – Fifth Edition (SB-5), or the Wechsler Intelligence Scales for Children/Wechsler Preschool and Primary Scale of Intelligence (WISC/WPPSI). From each assessment, verbal cognition or domain standard scores with a mean of 100 and a standard deviation of 15 were extracted. This score captures a child's cognitive abilities when verbal language abilities were included.

Language was calculated using one of the following: Clinical Evaluation of Language Fundamentals (CELF) (Wiig et al., 2013), Mullen Scales of Early Learning (MSEL), Bayley-4 (Bayley & Aylward, 2019), REEL-4 (Bzoch et al., 2020), Comprehensive Assessment of Spoken Language-Second Edition (CASL) (Carrow-Woolfolk, 2017), or the Preschool Language Scales (PLS). From each assessment, core language or communication standard scores with a mean of 100 and a standard deviation of 15 were extracted. This score captures a child's verbal language abilities within expressive and receptive domains. For the MSEL, T-scores were rescaled to standard score range for comparison (see above).

Adaptive behavior was calculated using one of the following: Vineland Adaptive Behavior Scales (Vineland-3), Adaptive Behavior Assessment System (ABAS) (Harrison & Oakland, 2015), Developmental Assessment of Young Children (DAYC) (Voress & Maddox, 2013), or the Battelle Developmental Inventory (BDI) (Newborg, 2005). From each assessment, overall adaptive behavior standard scores with a mean of 100 and a standard deviation of 15 were extracted. This score captures a child's overall life skills within domains such as daily living, communication, and socialization.

**Table S1: Harmonization of measures across several developmental and behavioral measures produces 4 domains for use in further analysis: nonverbal cognition, verbal cognition, verbal language, and adaptive behavior. "Score" indicates the item name used for the standard score in each domain; "Subcategories and Calculation" indicates the subscales that contributed to determining the score.**

| | Score | Subcategories & Calculation | n-size | |
|---|---|---|---|---|
| Nonverbal Cognition | | | TD | ASD |

| | | | | | |
|---|---|---|---|---|---|
| **Mullen Scales of Early Learning (MSEL)** | Nonverbal Domain (NVD) | Visual Reception T-Score | Fine Motor T-Score | 30 | 39 |
| **Differential Abilities Scale 2 (DAS-2)** | Nonverbal IQ (NVIQ) | School Age: Matrices, Sequential and Quantitative Reasoning | | | 4 |
| | | Early Years: Picture Similarities, Matrices | | | 2 |
| | | Unknown | | | 3 |
| **Wechsler Intelligence Scale for Children, Fifth Edition (WISC-V)** | Nonverbal Index (NVI) | Visual Spatial Index Standard Score (Block Design, Visual Puzzles) | Fluid Reasoning Index Standard Score (Matrix Reasoning, Figure Weights) | | 7 |
| **Sanford Binet 5 (SB-5)** | Nonverbal IQ (NVIQ) | Nonverbal: Fluid Reasoning, Knowledge, Quantitative Reasoning, Visual Spatial Processing, Working Memory | | | 1 |
| Verbal Cognition | | | | | |
| **Differential Abilities Scale 2 (DAS-2)** | Verbal IQ (VIQ) | School Age: Verbal Similarities, Word Definitions | | | 3 |
| | | Early Years: Verbal Comprehension, Naming Vocabulary | | | 2 |
| | | Unknown | | | 3 |
| **Wechsler Preschool and Primary Scale of Intelligence (WISC-V)** | Verbal Comprehension Index (VCI) | Similarities, Vocabulary, Information, Comprehension | | | 7 |
| **Wechsler Preschool and Primary Scale of Intelligence (WPPSI-V)** | Verbal Comprehension Index (VCI) | Information, Similarities | | | 1 |
| **Sanford Binet 5 (SB-5)** | Verbal IQ (VIQ) | Verbal: Fluid Reasoning, Knowledge, Quantitative Reasoning, Visual Spatial Processing, Working Memory | | | 1 |
| Verbal Language | | | | | |
| **Preschool Language Scales (PLS)** | Total Language Score | Expressive Language Standard Score | Receptive Language Standard Score | 28 | 33 |
| **Bayley-4** | Language | Receptive Communication (RC) | Expressive Communication (EC) | | 1 |
| **Mullen Scales of Early Learning (MSEL)** | Total Language Score | Expressive Language T-Score | Receptive Language T-Score | | 2 |

| | | | | | | |
|---|---|---|---|---|---|---|
| **Comprehensive Assessment of Spoken Language (CASL)** | General Language Ability Index | Lexical/Semantic, Syntactic, Supralinguistic, and Pragmatic language categories | | | | 3 |
| **Receptive-Expressive Emergent Language Test (REEL-4)** | Language Ability composite | Receptive Language | Expressive Language | | | 1 |
| **Clinical Evaluation of Language Fundamentals (CELF)** | Core Language Score | Receptive Language Index, Expressive Language Index, Language Content Index, Language Structure Index, Language Memory Index score | | | | 1 |
| Adaptive Behavior | | | | | | |
| **Vineland Adaptive Behavior Scales (Vineland-3)** | Adaptive Behavior Composite | Communication (Receptive, Expressive, and Written Language* Domains) | Daily Living (Personal, Domestic, and Community categories) | Socialization (Interpersonal Relationships, Play and Leisure, and Coping Skills) | 28 | 42 |
| **Adaptive Behavior Assessment System (ABAS)** | General Adaptive Composite | Conceptual (Communication, Functional Preacademics or Academics, Self-Direction) | Social (Leisure, Social) | Practical (Community Use, Home Living, Self-Care, Health and Safety) | | 4 |

### 2. PCA-based Electrode Selection

Conventional VEP analyses typically quantify responses at the occipital electrodes O1, Oz, and O2 (Šuštar Habjan et al., 2025). However, the use of a fixed electrode set may be suboptimal in developmental and clinical populations for both practical and neurophysiological reasons. Children with autism, as well as younger TD children, may have greater difficulty remaining still during sensor placement and recording. Although standardized procedures were used to maintain consistent net positioning, participant movement or pulling on the net could introduce variability in electrode positioning. In addition, the spatial distribution of VEP responses may vary across individuals and groups; for example, hemispheric asymmetries in visual processing have been reported in autism (Pei et al., 2014). Consequently, using the same fixed electrodes for all participants could result in suboptimal sampling of individual VEP responses and potentially contribute to apparent group differences.

To account for this variability, we implemented a data-driven procedure to select electrodes individually for each participant. We first defined a candidate set of electrodes over posterior occipital regions (Figure S2). Spatial principal component analysis (PCA) was then applied to detrended, channel-wise, trial-averaged event-related potentials within this candidate region to identify the dominant spatial patterns underlying each participant's VEP response. PCA components were ranked according to explained variance, and components cumulatively accounting for 80% of the total variance were retained. For each electrode, a channel weight was calculated as a variance-weighted combination of its loadings across the retained components, yielding a spatial profile representing the electrode's contribution to the dominant VEP patterns. The three electrodes with the highest weights were selected for subsequent VEP analyses. Importantly, PCA was used solely to identify electrodes and was not used to derive the VEP waveform itself; all subsequent VEP measures were calculated from the original EEG signals at the selected electrodes.

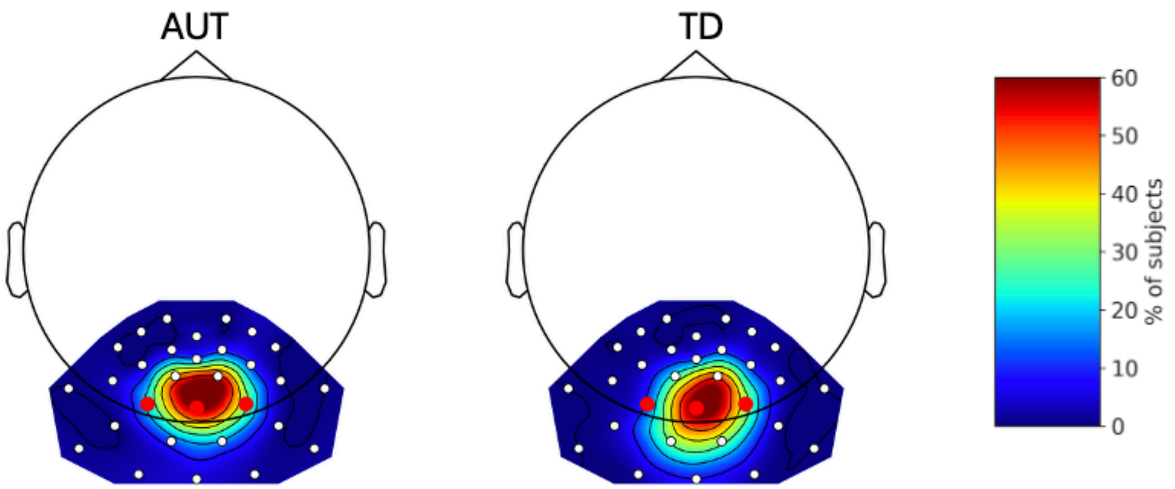


**Figure S2. Spatial distribution of electrodes selected for VEP analysis.** Topographic maps show the percentage of participants for whom each electrode was selected for quantification of the visual evoked potential (VEP) in the autism (AUT) and typically developing (TD) groups. Electrode selection was performed individually for each participant based on the magnitude of the VEP response within the predefined posterior electrode region. Warmer colors indicate electrodes selected in a greater proportion of participants. White dots indicate candidate electrode locations. Red dots indicate electrodes often used in conventional VEP analysis, O1, Oz, and O2.

To evaluate this individualized approach, we compared VEPs obtained from the PCA-selected electrodes with those obtained using the conventional O1, Oz, and O2 electrodes. Both approaches showed similar group-level patterns, while the individualized approach yielded larger VEP responses (Figure S3). The spatial distributions of selected electrodes were also similar between AUT and TD participants (Figure S2), providing no indication that systematic group differences in electrode selection accounted for the observed VEP findings.

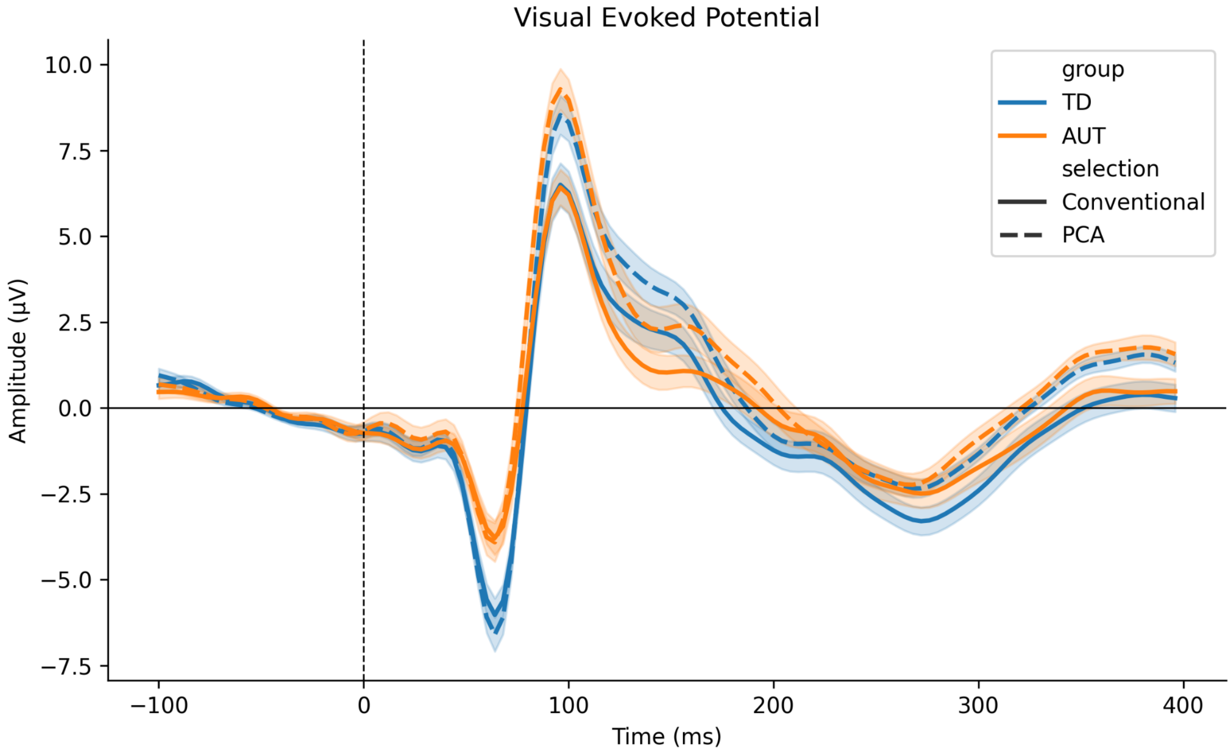


**Figure S3. Comparison of VEP waveforms derived using individualized and conventional electrode selection.** Grand-average visual evoked potential (VEP) waveforms are shown for autistic (AUT; orange) and typically developing (TD; blue) participants using individually PCA-selected electrodes (solid lines) and the conventional O1, Oz, and O2 electrodes (dashed lines). Shaded regions indicate the standard error of the mean. Both electrode-selection approaches yielded similar group-level waveform patterns, while the individualized PCA-based approach produced larger VEP responses. The vertical dashed line indicates stimulus onset (0 ms).

**Table S2. Associations between VEP and resting EEG measures**

| Measures | Group-adjusted Model | | | | Pooled Model (unadjusted for group) | | | |
|---|---|---|---|---|---|---|---|---|
| | Periodic Alpha | | Aperiodic Exponent | | Periodic Alpha | | Aperiodic Exponent | |
| | Beta | p | Beta | p | Beta | p | Beta | p |
| N1 Amplitude | -5.65 | 0.096 | 0.61 | 0.574 | **-6.84** | **0.042** | 0.27 | 0.805 |
| N1-P1 Amplitude | **12.62** | **0.007** | 1.31 | 0.386 | **13.59** | **0.003** | 1.61 | 0.281 |
| ITPC | **0.18** | **0.022** | 0.04 | 0.108 | **0.22** | **0.005** | **0.05** | **0.046** |

**Table S3. Association between VEP and developmental/behavioral measures within the AUT cohort**

| | **N1 amplitude** | **N1-P1 amplitude** | **ITPC** |
|---|---|---|---|
| **Nonverbal development (n=55)** | b=0.114, p=0.8039 | b=0.414, p=0.1851 | **b=48.6, p=0.0354** |
| **Verbal Cognition (n=17)** | b=-1.03, p=0.4001 | b=0.787, p=0.2759 | **b=74.5, p=0.0473** |
| **Language (n=45)** | b=-0.47, p=0.4134 | b=0.384, p=0.3561 | **b=66.6, p=0.0295** |
| **Adaptive Behavior (n=46)** | b=-0.313, p=0.3800 | b=-0.0967, p=0.7015 | b=5.81, p=0.7812 |
| **Social Responsiveness Scale (n=86)** | b=0.204, p=0.3292 | b=0.0202, p=0.8886 | b=-4.14, p=0.6656 |
